# Surveillance of bacterial and fungal infections in hospitalised COVID-19 patients: an ISARIC/WHO CCP-UK and UKHSA linkage study

**DOI:** 10.64898/2026.10.07.26363238

**Authors:** Chloe Davison, Emily Mason, Natalie King, Georgia Beckett-Hill, Jocelyn Elmes, Lance Turtle, Russell Hope, Cameron J. Fairfield, Thomas M. Drake, Malcolm G Semple, J. Kenneth Baillie, Colin S. Brown, Antonia Ho, Sarah M. Gerver

## Abstract

**Objectives:** To characterise COVID-19 inpatients with and without bacterial/fungal preceding, co- and secondary infections, and describe the distribution of organisms associated with COVID-19 and antibiotic use.

**Methods:** We linked ISARIC/WHO CCP-UK data from hospitals in England (admissions 01/01/2020-18/10/2021) with UK Health Security Agency infections datasets of COVID-19, and bacterial- and fungal-positive specimens using a multi-step hierarchical algorithm. We retained those aged ≥18 years and categorised COVID-19-associated infection as preceding (2-28 days prior to SARS-CoV-2 PCR positive date), co-infection (±1 day) or secondary (2-28 days after).

**Results:** During the study period, there were 173,947 hospitalised episodes of COVID-19; 5.8% (n=10,017) with a bacterial or fungal preceding, co- or secondary infection. Preceding and co-infections were rare (preceding: 0.7%; co-infection: 1.5%) while secondary infections were more common (3.5%). *Escherichia coli* (29.6%) and *Staphylococcus aureus* (15.2%) were the most identified organisms in blood specimens, while *S. aureus* (22.9%) and *Pseudomonas aeruginosa* (20.4%) were most common in lower respiratory tract specimens. The 28-day case fatality rate (CFR) remained static (36%) in those with secondary infection, whereas it decreased between early 2020 and mid-2021 in the other infection groups (37% to 27%). Of those with no positive microbiology, 72.8% received antibiotics, though this decreased between January-June 2020 (77.6%) and April-October 2021 (62.6%).

**Conclusion:** Bacterial and fungal infections complicated approximately 1-in-17 hospitalised episodes of COVID-19, with CFR of ≥25%. Antibiotic prescribing was common in those without firm evidence of bacterial infection. We recommend adhering to NICE guidance which discourages empirical antibiotic use for hospitalised COVID-19 patients.

## Introduction

The Coronavirus Disease 2019 (COVID-19) pandemic, caused by Severe Acute Respiratory Syndrome Coronavirus 2 (SARS-CoV-2), posed unprecedented challenges to healthcare systems worldwide [1]. Understanding infections temporally associated with COVID-19, including their timing and consequences, is essential for evaluating their impact on patient outcomes. Distinguishing co-infections from secondary infections is particularly important, as they have distinct implications for patient management, antimicrobial stewardship, and stockpiling for medical countermeasure preparedness.

In contrast to pandemic and seasonal influenza, where the prevalence of bacterial co-infections is ∼17-23% [2–4], evidence from the first COVID-19 wave suggested that bacterial co- and secondary infections were relatively uncommon. National surveillance data from England (January-June 2020) reported that 1.0% (95% CI: 0.97–1.06) of individuals with COVID-19 in primary and secondary care had a bacterial or fungal co- or secondary infection, including bloodstream, lower respiratory tract (LRT) and *Clostridiodes difficile* infections (CDI) [5]. However, higher estimates have been reported in hospitalised populations. A 2020 meta-analysis by Langford *et al.* found pooled estimate of 3.5% (95% CI: 0.4-6.7) for bacterial co-infection and 14.3% (95% CI: 9.6-18.9) for secondary infection in predominantly hospitalised patients [6]. This analysis included bloodstream, LRT and urinary specimens, and did not specify end of follow-up for secondary infections.

Patients with COVID-19 and concomitant infections experience worse outcomes than those with SARS-CoV-2 infection alone [7, 8], including longer length of stay (LoS), increased in-hospital mortality and higher case fatality rate (CFR) [5, 9–11]. Fungal co-infections have also contributed substantially to morbidity, exemplified by outbreaks of COVID-19-associated mucormycosis in India [5, 12–14].

Empirical antimicrobial prescribing in COVID-19-associated infections, particularly in the absence of microbiology confirmation, risks unnecessary adverse effects and may exacerbate antimicrobial resistance (AMR), a leading cause of mortality globally [15]. In England, overall antibiotic prescribing in secondary care decreased over 2020 compared with pre-pandemic levels; however, there was a marked increase in broad-spectrum antibiotic use per patient in April 2020 [16]. A large UK prospective cohort study estimated that 85% of hospitalised COVID-19 patients received antimicrobials during admission [17]. Furthermore, increases in COVID-19-related hospital admissions have been associated with increased AMR [18, 19]. These findings highlight the need to better characterise the prevalence and aetiology of COVID-19-associated infections, in addition to antimicrobial use to understand appropriateness of prescribing, inform management guidelines, and reduce the risk of worsening AMR.

In this study, we combined data from a prospective cohort of hospitalised COVID-19 patients in England with UK Health Security Agency (UKHSA) surveillance datasets to: (i) characterise the epidemiology and aetiology of COVID-19 associated infections; (ii) understand patterns of antibiotic use in those with and without bacterial/fungal infections across three waves of SARS-CoV-2 infections to inform clinical guidelines and stewardship strategies.

## Methods

The International Severe Acute Respiratory and emerging Infection Consortium (ISARIC) / World Health Organisation (WHO) Clinical Characterisation Protocol UK (CCP-UK) study was a prospective cohort study conducted across acute care hospitals in Great Britain between January 2020 and February 2022 [20, 21]. People admitted to hospital with confirmed SARS-CoV-2 infection, or with a high likelihood of infection during the first wave, were eligible for inclusion. Patients were followed up until hospital discharge or death.

SARS-CoV-2 positive PCR tests, and positive bacterial and fungal blood and LRT specimens were extracted from UKHSA’s Second-Generation Surveillance system (SGSS) [22]. Bacterial infections comprised blood and LRT specimens, and fungal infections included blood specimens for *Candida spp.* and respiratory specimens for *Aspergillus spp. Haemophilus influenzae* and *Streptococcus pneumoniae* data include only invasive infections identified in sterile sites (counted under blood specimens for this analysis). CDI data were extracted from UKHSA’s healthcare-associated infections (HCAI) data capture system (DCS).

### Data linkage and participant definition

UKHSA COVID-19 records for England (specimen dates 1^st^ January 2020 to 31^st^ December 2021) were linked to CCP-UK participants admitted in England between 1^st^ January 2020 and 18^th^ October 2021 (final admission date) using a multi-stage linkage algorithm (Supplementary Table S1). Individuals under 18 years old at admission were excluded. Unlinked UKHSA records were excluded to prioritise the CCP-UK case definition, which was not solely reliant on PCR confirmation. UKHSA COVID-19 records and CCP-UK data included date of birth, home postcode, ethnicity and sex. Additional fields in CCP-UK data included: comorbidities, ventilation use, hospital code, antibiotic use and, date of admission and discharge from hospital and intensive care unit (ICU).

These data were independently linked with microbiology data from SGSS and HCAI DCS (specimen dates between 1^st^ January 2020 and 14^th^ November 2021), allowing up to 27 days follow-up after final CCP-UK admission. Linkage used a multi-stage hierarchical algorithm (Supplementary Table S2). Infections occurring within 28 days of hospital admission were classified as COVID-19-associated. Patients without linked microbiological evidence of infection (within ±27 days of admission), together with unlinked CCP-UK records, formed the comparator group. Infection episodes were defined by organism and guided by infectious disease experts (COVID-19: rolling 90-day episodes; CDI: static 28-day; all others: site-specific rolling 14-day) [5].

### Data analysis

Infections were categorised according to timing relative to hospital admission. Preceding infections were defined as those occurring 2-27 days before CCP-UK date of study admission. Date of study entry was utilised rather than SARS-CoV-2 specimen date as this more accurately determined onset by relating to symptoms, rather than the confirmatory laboratory test. Co-infections were infections with a specimen date between 1 day before and 1 day after admission. Secondary infections were those with a specimen date 2-27 days after admission. *S. pneumoniae* and *H. influenzae* used adjusted definitions due to expert advice; co-infections were 2 days either side of admission and preceding and secondary infections from 3-27 days either side. COVID-19-associated infection status was mutually exclusive within COVID-19 episode, prioritised by earliest specimen date of linked infection. Specimen type categories were not mutually exclusive per episode and assigned based on the combination of blood, LRT and CDI positive specimens.

COVID-19 wave definition: wave 1 (W1), 1^st^ January 2020 – 28^th^ June 2020; wave 2 (W2), 29^th^ June 2020 – 26^th^ April 2021; wave 3 (W3), 27^th^ April 2021 – 18^th^ October 2021. Hospital and ICU LoS were calculated from admission and discharge dates. Ethnicity was grouped using the CHIME methodology [23] and index of multiple deprivation (IMD) was derived from patient postcode and categorised into quintiles. CFR was calculated including patients where death occurred within 28 days of SARS-CoV-2 PCR positivity.

Data linkage and analysis was carried out in Stata 17. CCP-UK data cleaning was carried out in R (Version 4.2.2).

## Results

### Characteristics of hospitalised COVID-19 patients

A total of 173,947 hospitalised COVID-19 episodes were included (Table 1). The majority of patients were ≥50 years old (77.3%, n=134,521/173,947), male (54.5%, n=94,847/173,947), and in the White ethnic group (68.3%, n=118,750/173,947). The highest regional representation was from the North West of England (18.9%, n=32,864/173,947) and 27.1% resided in the most deprived IMD quintile (n=47,196/173,947). Diabetes was the most common comorbidity (21.1%, n=36,754/173,947), followed by chronic pulmonary disease (CPD; 14.5%, n=25,184/173,947) and immunocompromising conditions (11.4%, n=19,752/173,947).

**Table 1:** Characteristics of CCP-UK patients with COVID-19 and with/without a preceding, co-, and secondary infection with a hospital admission across the COVID-19 waves. Wave 1 (W1): 1^st^ January 2020 to 28^th^ June 2020. This differs from the UK definitions of wave 1 starting on 30^th^ January 2020. Wave 2 (W2): 29^th^ June 2020 to 26^th^ April 2021. Wave 3 (W3): 27^th^ April 2021 to study end (last CCP-UK admitted patient 18^th^ October 2021). Values under 8 and their percentages censored with a ^ and other values in the same category rounded to the nearest 5.

|  | Total<br>(n=173,947) |  | COVID-19 patients without preceding/<br>co-/secondary infection |  |  |  |  |  | COVID-19 patients with<br>preceding/co-infection |  |  |  |  |  | COVID-19 patients with<br>secondary infection |  |  |  |  |  |
| --- | --- | --- | --- | --- | --- | --- | --- | --- | --- | --- | --- | --- | --- | --- | --- | --- | --- | --- | --- | --- |
|  |  |  | W1<br>(n=56,815) |  | W2<br>(n=85,400) |  | W3<br>(n=21,715) |  | W1<br>(n=1,504) |  | W2<br>(n=2,056) |  | W3<br>(n=303) |  | W1<br>(n=1,767) |  | W2<br>(n=3,703) |  | W3<br>(n=684) |  |
|  | N | % | N | % | N | % | N | % | N | % | N | % | N | % | N | % | N | % | N | % |
| Age (years) |  |  |  |  |  |  |  |  |  |  |  |  |  |  |  |  |  |  |  |  |
| <50 | 32,959 | 18.9 | 7,211 | 12.7 | 14,942 | 17.5 | 9,383 | 43.2 | 103 | 6.8 | 188 | 9.1 | 60 | 19.8 | 276 | 15.6 | 555 | 15.0 | 240 | 35.1 |
| 50 to 69 | 46,847 | 26.9 | 14,062 | 24.8 | 23,733 | 27.8 | 5,329 | 24.5 | 323 | 21.5 | 492 | 23.9 | 80 | 26.4 | 816 | 46.2 | 1,735 | 46.9 | 280 | 40.9 |
| 70 to 79 | 34,795 | 20.0 | 12,170 | 21.4 | 17,374 | 20.3 | 3,012 | 13.9 | 394 | 26.2 | 543 | 26.4 | 70 | 23.1 | 320 | 18.1 | 814 | 22.0 | 100 | 14.6 |
| ≥80 | 52,879 | 30.4 | 20,748 | 36.5 | 26,329 | 30.8 | 3,456 | 15.9 | 620 | 41.2 | 794 | 38.6 | 90 | 29.7 | 261 | 14.8 | 513 | 13.9 | 65 | 9.5 |
| Unknown | 6,467 | 3.7 | 2,624 | 4.6 | 3,022 | 3.5 | 535 | 2.5 | 64 | 4.3 | 39 | 1.9 | ^ | ^ | 94 | 5.3 | 86 | 2.3 | ^ | ^ |
| Sex |  |  |  |  |  |  |  |  |  |  |  |  |  |  |  |  |  |  |  |  |
| Female | 78,818 | 45.3 | 25,098 | 44.2 | 39,950 | 46.8 | 10,247 | 47.2 | 600 | 39.9 | 825 | 40.1 | 120 | 39.6 | 560 | 31.7 | 1,185 | 32.0 | 233 | 34.1 |
| Male | 94,842 | 54.5 | 31,600 | 55.6 | 45,325 | 53.1 | 11,437 | 52.7 | 900 | 59.8 | 1,230 | 59.8 | 183 | 60.4 | 1,205 | 68.2 | 2,515 | 67.9 | 451 | 65.9 |
| Unknown | 287 | 0.2 | 117 | 0.2 | 125 | 0.1 | 31 | 0.1 | ^ | ^ | ^ | ^ | 0 | - | ^ | ^ | ^ | ^ | 0 | - |
| Region |  |  |  |  |  |  |  |  |  |  |  |  |  |  |  |  |  |  |  |  |
| East Midlands | 16,498 | 9.5 | 4,985 | 8.8 | 9,056 | 10.6 | 1,527 | 7.0 | 130 | 8.6 | 242 | 11.8 | 25 | 8.3 | 112 | 6.3 | 376 | 10.2 | 47 | 6.9 |
| East of England | 21,118 | 12.1 | 6,726 | 11.8 | 11,126 | 13.0 | 1998 | 9.2 | 195 | 13.0 | 322 | 15.7 | 29 | 9.6 | 256 | 14.5 | 397 | 10.7 | 68 | 9.9 |
| London | 14,487 | 8.3 | 6,518 | 11.5 | 5,436 | 6.4 | 1,555 | 7.2 | 135 | 9.0 | 109 | 5.3 | 21 | 6.9 | 279 | 15.8 | 376 | 10.2 | 60 | 8.8 |
| North East | 7,804 | 4.5 | 2,627 | 4.6 | 3,436 | 4.0 | 1,393 | 6.4 | 60 | 4.0 | 65 | 3.2 | 13 | 4.3 | 48 | 2.7 | 127 | 3.4 | 35 | 5.1 |
| North West | 32,864 | 18.9 | 10,485 | 18.5 | 16,052 | 18.8 | 4,289 | 19.8 | 340 | 22.6 | 412 | 20.0 | 51 | 16.8 | 377 | 21.3 | 677 | 18.3 | 179 | 26.2 |
| South East | 23,993 | 13.8 | 7,495 | 13.2 | 12,054 | 14.1 | 2,840 | 13.1 | 245 | 16.3 | 301 | 14.6 | 41 | 13.5 | 307 | 17.4 | 636 | 17.2 | 74 | 10.8 |
| South West | 11,766 | 6.8 | 3,190 | 5.6 | 6,043 | 7.1 | 1,887 | 8.7 | 90 | 6.0 | 137 | 6.7 | 28 | 9.2 | 86 | 4.9 | 254 | 6.9 | 52 | 7.6 |
| West Midlands | 22,807 | 13.1 | 7,996 | 14.1 | 10,895 | 12.8 | 2,557 | 11.8 | 200 | 13.3 | 284 | 13.8 | 42 | 13.9 | 189 | 10.7 | 560 | 15.1 | 85 | 12.4 |
| Yorkshire and Humber | 18,178 | 10.5 | 5,160 | 9.1 | 9,074 | 10.6 | 3,099 | 14.3 | 110 | 7.3 | 184 | 8.9 | 53 | 17.5 | 113 | 6.4 | 300 | 8.1 | 84 | 12.3 |
| Unknown | 4,432 | 2.5 | 1,633 | 2.9 | 2,228 | 2.6 | 570 | 2.6 | ^ | ^ | 0 | - | 0 | - | 0 | - | 0 | - | 0 | - |
| Ethnicity |  |  |  |  |  |  |  |  |  |  |  |  |  |  |  |  |  |  |  |  |
| White | 118,750 | 68.3 | 38,027 | 66.9 | 59,826 | 70.1 | 13,844 | 63.8 | 1,179 | 78.4 | 1,663 | 80.9 | 225 | 74.3 | 1,085 | 61.4 | 2,470 | 66.7 | 433 | 63.3 |
| Black | 6,024 | 3.5 | 2,295 | 4.0 | 2,425 | 2.8 | 899 | 4.1 | 48 | 3.2 | 48 | 2.3 | 15 | 5.0 | 130 | 7.4 | 132 | 3.6 | 33 | 4.8 |
| Asian | 12,988 | 7.5 | 3,648 | 6.4 | 6,660 | 7.8 | 1,772 | 8.2 | 59 | 3.9 | 100 | 4.9 | 25 | 8.3 | 193 | 10.9 | 454 | 12.3 | 79 | 11.5 |
| Mixed/Other | 3,665 | 2.1 | 1,370 | 2.4 | 1,496 | 1.8 | 578 | 2.7 | 26 | 1.7 | 28 | 1.4 | 10 | 3.3 | 68 | 3.8 | 76 | 2.1 | 15 | 2.2 |
| Unknown | 32,520 | 18.7 | 11,475 | 20.2 | 14,993 | 17.6 | 4,622 | 21.3 | 192 | 12.8 | 217 | 10.6 | 35 | 11.6 | 291 | 16.5 | 571 | 15.4 | 124 | 18.1 |
| IMD quintile |  |  |  |  |  |  |  |  |  |  |  |  |  |  |  |  |  |  |  |  |
| 1 (least deprived) | 24,723 | 14.2 | 8,429 | 14.8 | 12,336 | 14.4 | 2,539 | 11.7 | 230 | 15.3 | 314 | 15.3 | 33 | 10.9 | 280 | 15.8 | 497 | 13.4 | 67 | 9.8 |
| 2 | 29,062 | 16.7 | 9,801 | 17.3 | 14,344 | 16.8 | 3,267 | 15.0 | 275 | 18.3 | 349 | 17.0 | 53 | 17.5 | 313 | 17.7 | 575 | 15.5 | 86 | 12.6 |
| 3 | 31,898 | 18.3 | 10,544 | 18.6 | 15,701 | 18.4 | 3,721 | 17.1 | 280 | 18.6 | 419 | 20.4 | 59 | 19.5 | 324 | 18.3 | 734 | 19.8 | 118 | 17.3 |
| 4 | 36,636 | 21.1 | 11,928 | 21.0 | 17,772 | 20.8 | 4,721 | 21.7 | 340 | 22.6 | 431 | 21.0 | 65 | 21.5 | 402 | 22.8 | 811 | 21.9 | 165 | 24.1 |
| 5 (most deprived) | 47,196 | 27.1 | 14,480 | 25.5 | 23,019 | 27.0 | 6,897 | 31.8 | 380 | 25.3 | 543 | 26.4 | 93 | 30.7 | 448 | 25.4 | 1,086 | 29.3 | 248 | 36.3 |
| Unknown | 4,432 | 2.5 | 1,633 | 2.9 | 2,228 | 2.6 | 570 | 2.6 | ^ | ^ | 0 | - | 0 | - | 0 | - | 0 | - | 0 | - |
| **Comorbidities\*** |  |  |  |  |  |  |  |  |  |  |  |  |  |  |  |  |  |  |  |  |
| Chronic pulmonary disease | 25,184 | 14.5 | 9,482 | 16.7 | 11,892 | 13.9 | 2,205 | 10.2 | 295 | 19.6 | 401 | 19.5 | 60 | 19.8 | 239 | 13.5 | 529 | 14.3 | 81 | 11.8 |
| Immunocompromise | 19,752 | 11.4 | 6,453 | 11.4 | 9,589 | 11.2 | 2,153 | 9.9 | 247 | 16.4 | 349 | 17.0 | 55 | 18.2 | 229 | 13.0 | 566 | 15.3 | 111 | 16.2 |
| Diabetes | 36,754 | 21.1 | 9,959 | 17.5 | 20,534 | 24.0 | 3,774 | 17.4 | 329 | 21.9 | 617 | 30.0 | 73 | 24.1 | 315 | 17.8 | 998 | 27.0 | 155 | 22.7 |
| **Hospital outcomes\*** |  |  |  |  |  |  |  |  |  |  |  |  |  |  |  |  |  |  |  |  |
| Length of hospital stay** | 159,102 | 91.5 | 9 (4-17) |  | 9 (4-17) |  | 5 (3-10) |  | 14 (7-24) |  | 16 (9-26) |  | 10 (5-18) |  | 23 (13-39) |  | 19 (12-28) |  | 15 (9-23) |  |
| ICU admission | 25,085 | 14.4 | 6,229 | 11.0 | 11,426 | 13.4 | 2,753 | 12.7 | 205 | 13.6 | 357 | 17.4 | 79 | 26.1 | 1,122 | 63.5 | 2,467 | 66.7 | 447 | 65.4 |
| Length of ICU stay** | 20,277 | 11.7 | 7 (3-13) |  | 6 (3-10) |  | 5 (3-9) |  | 7 (4-15) |  | 9 (4-17) |  | 6 (4-10) |  | 20 (10-30) |  | 15 (8-25) |  | 13 (7-20) |  |
| Antibiotic use during admission*** | 127,976 | 73.6 | 44,121 | 77.6 | 61,618 | 72.2 | 13,607 | 62.6 | 1,252 | 84.1 | 1,773 | 87.1 | 250 | 82.8 | 1,475 | 85.9 | 3,243 | 89.0 | 568 | 85.9 |
| **Outcomes at ≤ 28 days** |  |  |  |  |  |  |  |  |  |  |  |  |  |  |  |  |  |  |  |  |
| Death | 40,070 | 23.0 | 17,220 | 30.3 | 17,280 | 20.2 | 2,000 | 9.2 | 580 | 38.6 | 690 | 33.6 | 81 | 26.7 | 570 | 32.3 | 1,429 | 38.6 | 222 | 32.5 |
| Discharged alive | 115,075 | 66.2 | 34,350 | 60.5 | 58,185 | 68.1 | 18,480 | 85.1 | 741 | 49.3 | 1,003 | 48.8 | 173 | 57.1 | 780 | 44.1 | 1,076 | 29.1 | 284 | 41.5 |
| Hospitalisation | 8,830 | 5.1 | 1,435 | 2.5 | 5,415 | 6.3 | 730 | 3.4 | 66 | 4.4 | 193 | 9.4 | 29 | 9.6 | 145 | 8.2 | 722 | 19.5 | 93 | 13.6 |
| Transfer to other facility/Palliative discharge | 9,970 | 5.7 | 3,810 | 6.7 | 4,520 | 5.3 | 505 | 2.3 | 117 | 7.8 | 170 | 8.3 | 20 | 6.6 | 270 | 15.3 | 476 | 12.9 | 85 | 12.4 |
| Unknown / Awaited | 10 | 0.0 | ^ | ^ | ^ | ^ | ^ | ^ | 0 | - | 0 | - | 0 | - | ^ | ^ | 0 | - | 0 | - |

\*Sub-categories within these categories are not mutually exclusive. \*\* median days (IQR). \*\*\* n values are different for this row due to excluding fungal specimens for antibiotic analysis: no COVID-19-associated bacterial infection W1 n=56,880, W2 n=85,481, and W3 n=21,739; bacterial preceding or co-infection W1 n=1488, W2 n=2,036, and W3 n=302; bacterial secondary infection W1 n=1,718, W2 n=3,642, and W3 n=661.

Bacterial or fungal COVID-19-associated infection was identified in 5.8% of hospitalised episodes (n=10,017/173,947; Table 2). The monthly percentage varied over time, peaking at 9.7% in Feb 2021 (n=103/1058; Figure 1). Preceding and co-infection were rare (0.7%, n=1,179/173,947 and 1.5%, n=2,684/173,947, respectively), while secondary infection was more common (3.5%, n=6,154/173,947).

**Figure 1:**
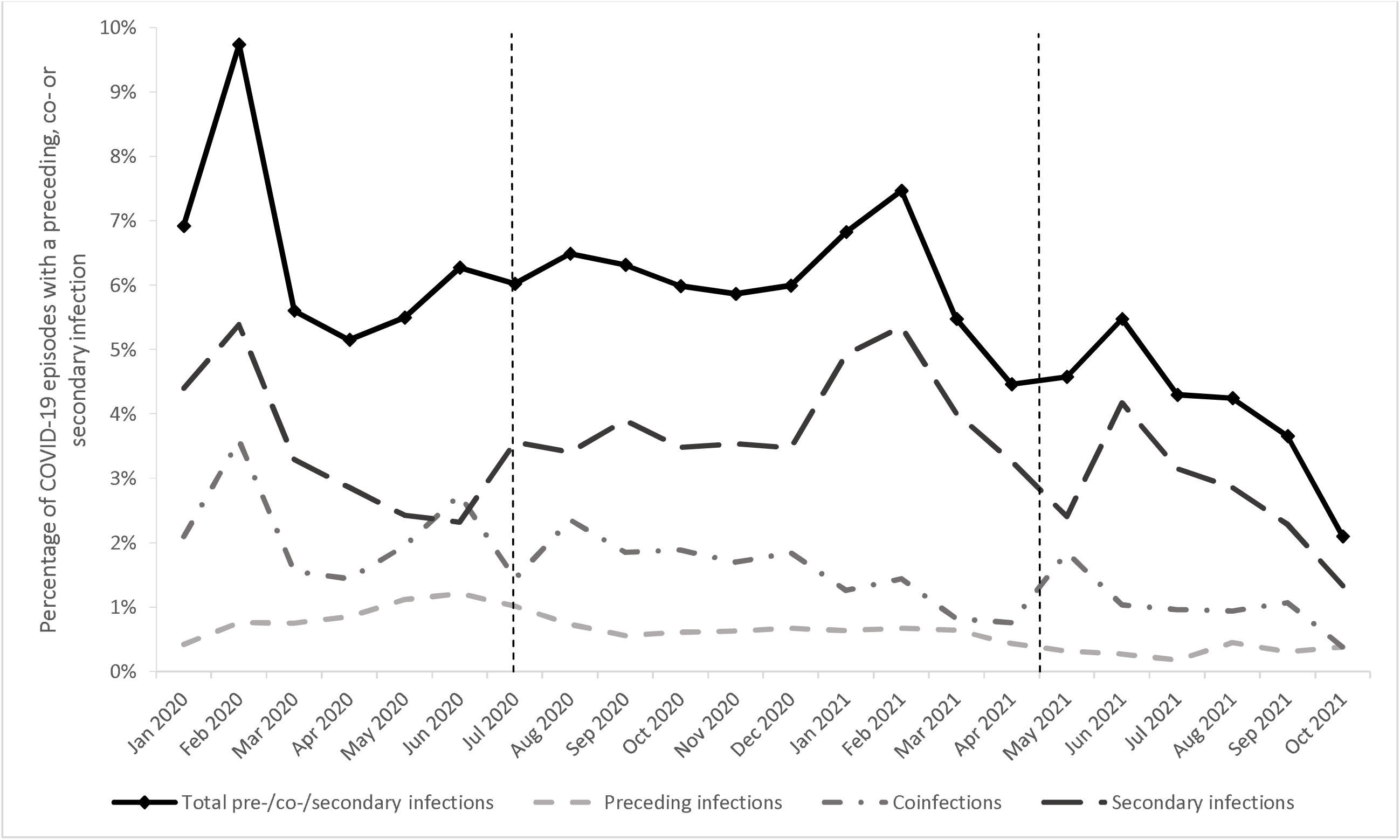
The percentage of preceding, co-, and secondary infections in CCP-UK patients with COVID-19 by date of hospital admission, within COVID-19 wave. Wave 1 (W1): 1^st^ January 2020 to 28^th^ June 2020. This differs from the UK definitions of wave 1 starting on 30^th^ January 2020. Wave 2 (W2): 29^th^ June 2020 to 26^th^ April 2021. Wave 3 (W3): 27^th^ April 2021 to study end (last CCP-UK admitted patient 18^th^ October 2021)

**Table 2:** The frequency of key bacterial/fungal infections in English CCP-UK patients with COVID-19 (N=173,947), by specimen type and timing of diagnosis. Data aren’t mutually exclusive, so a patient is included multiple times in this table apart from the ‘Any site’ row, in which case this refers to number of patients. Of note, 639 patients had both a BSI and a LRTI; 42 had a BSI and CDI; and, 17 had both a LRTI and CDI. 23 had a preceding infection and a secondary infection; and 48 had a co-infection and secondary infection. There were fewer than 10 patients with BSI, LRTI and CDI. Additionally, there were fewer than 10 patients with preceding and co-infection, or preceding, co- and secondary infection.

| Bacterial/ fungal infection by specimen type | COVID-19 patients with bacterial/ fungal infection |  | Timing of bacterial/fungal diagnosis in relation to COVID-19 diagnosis |  |  |  |  |  |  |  |  |
| --- | --- | --- | --- | --- | --- | --- | --- | --- | --- | --- | --- |
|  |  |  | Preceding infection |  |  | Co-infection |  |  | Secondary infection |  |  |
|  | n | % of COVID-19 patients | n | % infections by site | % of COVID-19 patients | n | % infections by site | % of COVID-19 patients | n | % infections by site | % of COVID-19 patients |
| Bloodstream infection | 5,273 | 3.0% | 750 | 14.2% | 0.4% | 2,244 | 42.6% | 1.3% | 2,279 | 43.2% | 1.3% |
| Lower respiratory infection | 4,913 | 2.8% | 406 | 8.3% | 0.2% | 420 | 8.5% | 0.2% | 4,087 | 83.2% | 2.3% |
| *Clostridioides difficile* infection | 541 | 0.3% | 86 | 15.9% | 0.0% | 122 | 22.6% | 0.1% | 333 | 61.6% | 0.2% |
| **Any site** | 10,017 | 5.8% | 1,179 | 11.8% | 0.7% | 2,684 | 26.8% | 1.5% | 6,154 | 61.4% | 3.5% |

Infection distribution differed by specimen type. Overall, 3.0% (n=5,273/173,947) of patients had bacteraemia or fungaemia, of which 42.6% (n=2,244/5,273) were co-infections and 43.2% (n=2,279/5,273) were secondary infections. Positive LRT samples were identified in 2.8% (n=4,913/173,947) of patients, predominantly secondary infections (83.2%, n=4,087/4,913). CDI were rare (0.3%, n=541/173,947), with 61.6% classified as secondary infection (n=333/541).

### Key organisms

Among patients with preceding/co-infection, the three most common causes of bacteraemia/fungaemia were consistent across waves (Table 3a): *Escherichia coli* (33.5-40.7% across waves, total n=1,044/2,994), *Staphylococcus aureus* (15.6-17.5%, n=514/2,994) and *Klebsiella pneumoniae* (7.6-8.5%, n=233/2,994).

**Table 3:** The most common causative organisms of preceding, co-, and secondary infections in (a) blood specimens and (b) lower respiratory specimens, in CCP-UK patients with COVID-19 by COVID-19 wave.

**a. Blood specimens**
|  | COVID-19 patients with bacterial/ fungal infection |  |  |  |  |  |  |  |  |
| --- | --- | --- | --- | --- | --- | --- | --- | --- | --- |
| Organisms in blood specimens | Wave 1 (n=1,888) |  |  | Wave 2 (n=3,002) |  |  | Wave 3 (n=383) |  |  |
|  | Rank | Count | % | Rank | Count | % | Rank | Count | % |
| *Escherichia coli* | 1 | 584 | 30.9 | 1 | 854 | 28.4 | 1 | 124 | 32.4 |
| *Staphylococcus aureus* | 2 | 279 | 14.8 | 2 | 463 | 15.4 | 2 | 59 | 15.4 |
| *Enterococcus faecium* | 3 | 182 | 9.6 | 3 | 308 | 10.3 | =4 | 23 | 6.0 |
| *Klebsiella pneumoniae* | 4 | 168 | 8.9 | 4 | 304 | 10.1 | 3 | 39 | 10.2 |
| *Enterococcus faecalis* | 5 | 89 | 4.7 | 6 | 136 | 4.5 | 7 | 22 | 5.7 |
|  | COVID-19 patients with preceding or co- bacterial/ fungal infection |  |  |  |  |  |  |  |  |
| Organisms in blood specimens | Wave 1 (n=1,203) |  |  | Wave 2 (n=1,592) |  |  | Wave 3 (n=199) |  |  |
|  | Rank | Count | % | Rank | Count | % | Rank | Count | % |
| *Escherichia coli* | 1 | 429 | 35.7 | 1 | 534 | 33.5 | 1 | 81 | 40.7 |
| *Staphylococcus aureus* | 2 | 211 | 17.5 | 2 | 272 | 17.1 | 2 | 31 | 15.6 |
| *Klebsiella pneumoniae* | 3 | 91 | 7.6 | 3 | 125 | 7.9 | 3 | 17 | 8.5 |
| *Proteus mirabilis* | 4 | 54 | 4.5 | 4 | 72 | 4.5 | =5 | ^ | ^ |
| *Enterococcus faecalis* | 5 | 47 | 3.9 | 7 | 56 | 3.5 | 4 | 11 | 5.5 |
|  | COVID-19 patients with secondary bacterial/ fungal infection |  |  |  |  |  |  |  |  |
| Organisms in blood specimens | Wave 1 (n=685) |  |  | Wave 2 (n=1,410) |  |  | Wave 3 (n=184) |  |  |
|  | Rank | Count | % | Rank | Count | % | Rank | Count | % |
| *Escherichia coli* | 1 | 155 | 22.6 | 1 | 320 | 22.7 | 1 | 43 | 23.4 |
| *Enterococcus faecium* | 2 | 149 | 21.8 | 2 | 264 | 18.7 | 4 | 19 | 10.3 |
| *Staphylococcus aureus* | 4 | 68 | 9.9 | 3 | 191 | 13.5 | 2 | 28 | 15.2 |
| *Klebsiella pneumoniae* | 3 | 77 | 11.2 | 4 | 179 | 12.7 | 3 | 22 | 12.0 |
| *Pseudomonas aeruginosa* | 7 | 33 | 4.8 | 5 | 111 | 7.9 | 5 | 18 | 9.8 |

**b. Lower respiratory tract specimens**
|  | COVID-19 patients with bacterial/ fungal infection |  |  |  |  |  |  |  |  |
| --- | --- | --- | --- | --- | --- | --- | --- | --- | --- |
| Organisms | Wave 1 (n=1,360) |  |  | Wave 2 (n=2,926) |  |  | Wave 3 (n=627) |  |  |
|  | Rank | Count | % | Rank | Count | % | Rank | Count | % |
| *Staphylococcus aureus* | 2 | 247 | 18.2 | 1 | 697 | 23.8 | 1 | 183 | 29.2 |
| *Pseudomonas aeruginosa* | 1 | 265 | 19.5 | 2 | 632 | 21.6 | 2 | 107 | 17.1 |
| *Klebsiella pneumoniae* | 4 | 201 | 14.8 | 3 | 519 | 17.7 | 3 | 91 | 14.5 |
| *Escherichia coli* | 3 | 230 | 16.9 | 4 | 458 | 15.7 | 4 | 85* | 13.6 |
| *Enterobacter cloacae complex* | 6 | 88 | 6.5 | 5 | 272 | 9.3 | 5 | 62 | 9.9 |
|  | COVID-19 patients with preceding or co- bacterial/ fungal infection |  |  |  |  |  |  |  |  |
| Organisms | Wave 1 (n=279) |  |  | Wave 2 (n=442) |  |  | Wave 3 (n=105) |  |  |
|  | Rank | Count | % | Rank | Count | % | Rank | Count | % |
| *Staphylococcus aureus* | 1 | 72 | 25.8 | 1 | 120 | 27.1 | 1 | 36 | 34.3 |
| *Pseudomonas aeruginosa* | 2 | 57 | 20.4 | 2 | 101 | 22.9 | 2 | 20 | 19.0 |
| *Escherichia coli* | 3 | 37 | 13.3 | 3 | 53 | 12.0 | =7 | ^ | ^ |
| *Klebsiella pneumoniae* | 4 | 25 | 9.0 | 4 | 32 | 7.2 | 3 | 15 | 14.3 |
| *Serratia marcescens/ureilytica* | 5 | 20 | 7.2 | 5 | 26 | 5.9 | =4 | ^ | ^ |
|  | COVID-19 patients with secondary bacterial/ fungal infection |  |  |  |  |  |  |  |  |
| Organisms | Wave 1 (n=1,081) |  |  | Wave 2 (n=2,484) |  |  | Wave 3 (n=522) |  |  |
|  | Rank | Count | % | Rank | Count | % | Rank | Count | % |
| *Staphylococcus aureus* | 4 | 175 | 16.2 | 1 | 577 | 23.2 | 1 | 147 | 28.2 |
| *Pseudomonas aeruginosa* | 1 | 208 | 19.2 | 2 | 531 | 21.4 | 2 | 87 | 16.7 |
| *Klebsiella pneumoniae* | 3 | 176 | 16.3 | 3 | 487 | 19.6 | 4 | 76 | 14.6 |
| *Escherichia coli* | 2 | 191 | 17.7 | 4 | 405 | 16.3 | 3 | 80* | 15.3 |
| *Enterobacter cloacae complex* | 5 | 82 | 7.6 | 5 | 253 | 10.2 | 5 | 62 | 11.9 |

Wave 1 (W1): 1<sup>st</sup> January 2020 to 28<sup>th</sup> June 2020. This differs from the UK definitions of wave 1 starting on 30<sup>th</sup> January 2020. Wave 2 (W2): 29<sup>th</sup> June 2020 to 26<sup>th</sup> April 2021. Wave 3 (W3): 27<sup>th</sup> April 2021 to study end (last CCP-UK admitted patient 18<sup>th</sup> October 2021). = denotes joint ranking with another organism. Values under 8 and their percentages censored with a ^ and other values for the same species rounded to the nearest 5 (marked with a \*).

For secondary infections, the top four causative organisms of bacteraemia/fungaemia were stable overall, although their relative ranking varied by wave. *E. coli* was the most common (22.6-23.4% across waves, total n=518/2,279), followed by *Enterococcus faecium* (10.3-21.8%, n=432/2,279), *S. aureus* (9.9%-15.2%, n=287/2,279) and *K. pneumoniae* (11.2-12.7%, n=278/2,279). No cases of *Haemophilus influenzae* and fewer than 10 cases of *Streptococcus pneumoniae* invasive infections were identified.

In the lower respiratory tract, organisms in preceding or co-infections were similar between waves one and two but differed in wave three (Table 3b). The most common organism was *S. aureus* (25.8-34.3% across waves, total n=228/826) followed by *Pseudomonas aeruginosa* (19.0-22.9%, n=178/826). Among secondary LRT infections, the four most common organisms, *S. aureus* (16.2-28.2%, n=899/4,087), *P. aeruginosa* (16.7-21.4%, n=826/4,087), *K. pneumoniae* (14.6-19.6%, n=739/4,087) and *E. coli* (15.7-17.7%, n=678/4,087), were consistent across waves with variation in ranking.

### Antibiotic therapy

Antibiotics were prescribed in 85.6% (n=3,275/3,826) of episodes with bacterial preceding or co-infection, and 87.8% (n=5,286/6,021) with bacterial secondary infection, compared with 72.8% (n=119,415/164,100) in patients without documented COVID-19-associated bacterial infection (Table 1). Antibiotic use in patients with COVID-19-associated bacterial infections peaked in wave 2 (preceding/co-infection: 87.1%, n=1,773/2,036; secondary: 89.0%, n=3,243/3,642), with a modest decline in wave 3 (preceding/co-infection: 82.8%, n=250/302; secondary: 85.9%, n=568/661). Among patients without COVID-19-associated bacterial infection, antibiotic use decreased across waves (77.6%, n=44,121/56,880 to 62.6%, n=13,607/21,739) but remained high throughout.

Penicillin with beta-lactamase inhibitors were the most frequently used antibiotics. Over 40% of patients without COVID-19-associated bacterial infection received this class (42.3%, n=69,425/164,100; Figure 2a). In bacterial preceding/co-infection, 55.4% (n=2,121/3,826) received penicillin with beta-lactamase inhibitors, followed by penicillin excluding beta-lactamase inhibitors (27.1%, n=1,037/3,826) and other beta-lactams (25.6%, n=981/3,826; Figure 2b). In bacterial secondary infections, use of penicillin with beta-lactamase inhibitor was higher (64.6%, n=3,889/6,021), alongside macrolides, lincosamindes and streptogramins (39.9%, n=2,404/6,021), and other beta-lactams (35.6%, n=2,144/6,021; Figure 2c).

**Figure 2:**
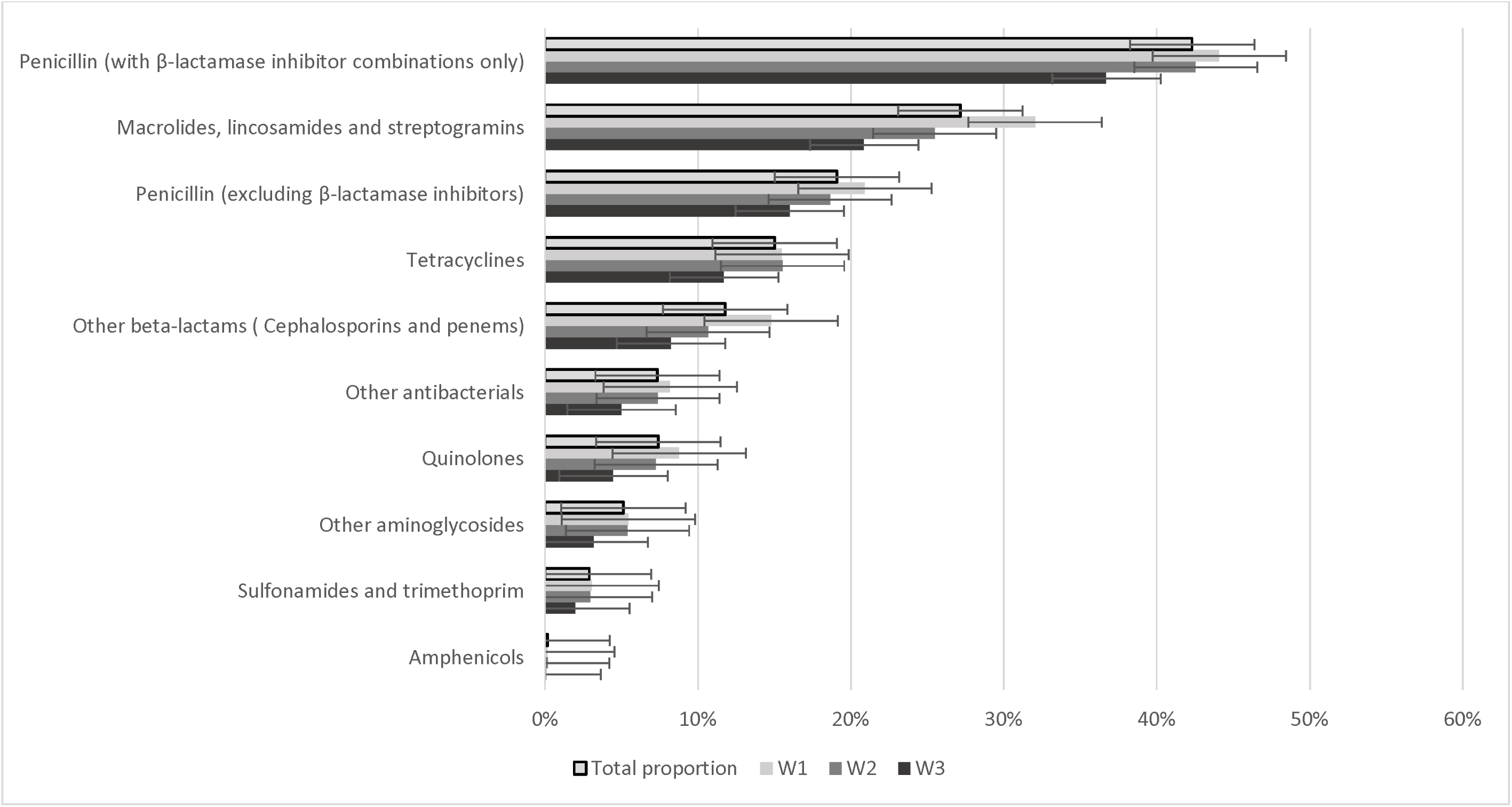
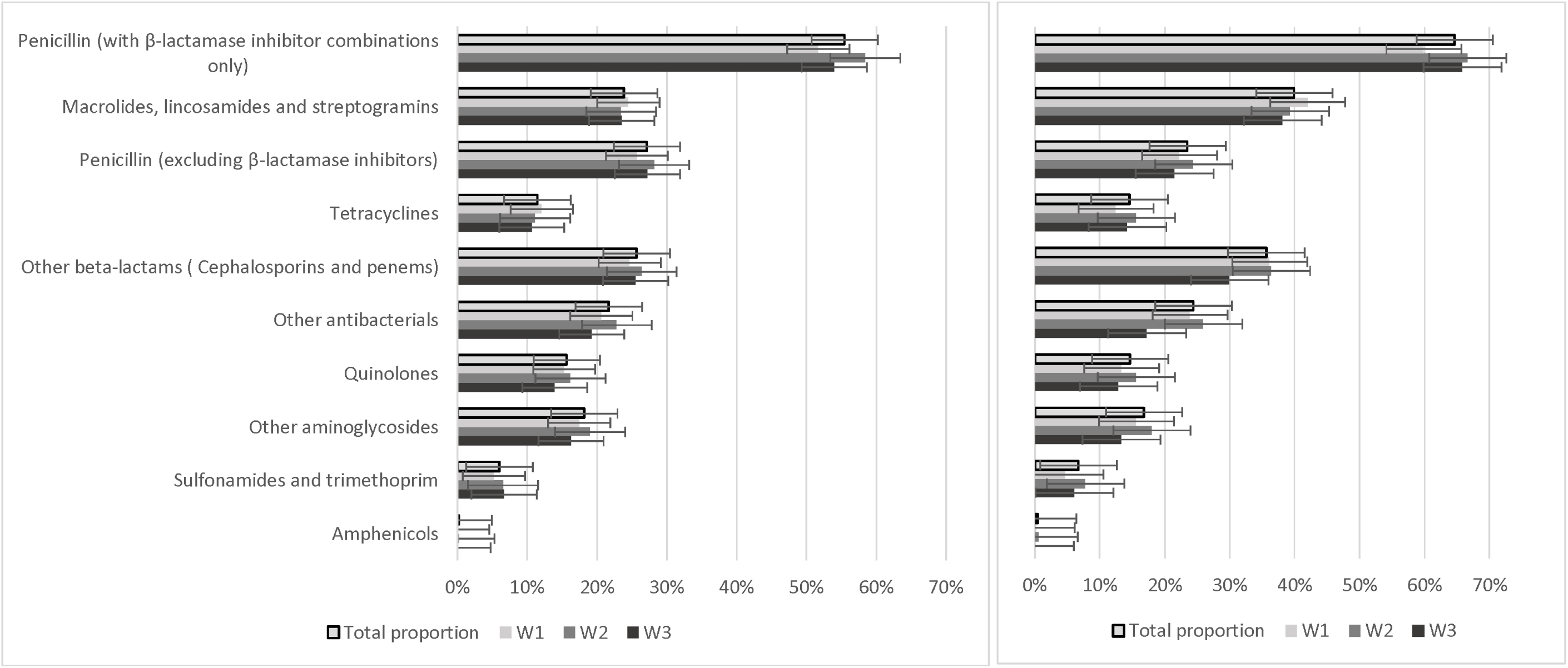
Antibiotics prescribed during hospital admission in CCP-UK patients: (a) inpatients with no COVID-19-associated bacterial infection, (b) those with COVID-19 and a bacterial preceding or co-infection, and (c) those with COVID-19 and a bacterial secondary infection, by hospital admission during COVID-19 waves. Wave 1 (W1): 1^st^ January 2020 to 28^th^ June 2020. This differs from the UK definitions of wave 1 starting on 30^th^ January 2020. Wave 2 (W2): 29^th^ June 2020 to 26^th^ April 2021. Wave 3 (W3): 27^th^ April 2021 to study end (last CCP-UK admitted patient 18^th^ October 2021). Please note, it is possible for a patient to have received one or more antibiotics, therefore a patient can be counted more than once.

### Outcomes

Median hospital LoS was highest among those with secondary infection, but decreased over time, from 23 days in wave 1 (interquartile range [IQR] 13-39 days) to 15 days (IQR 9-23 days) in wave 3 (Table 1).

Overall, 14.4% of hospitalised COVID-19 episodes required ICU admission (n=25,085/173,947) (Table 1). Patients with secondary infection had substantially higher ICU admission rates across the three waves (range across all waves: 63.5-66.7%, total n=4,036/6,154), compared to those without bacterial or fungal infection (11.0-13.4%, n=20,408/163,930) or with preceding/co-infection (13.6-26.1%, n=641/3,863). Among patients with secondary infection and ICU admission, 97.1% had a record of admission date (n=3,919/4,036). Of these, 95.2% developed secondary infection at least one day after ICU admission (n=3,729/3,919) (Figure 3).

**Figure 3:**
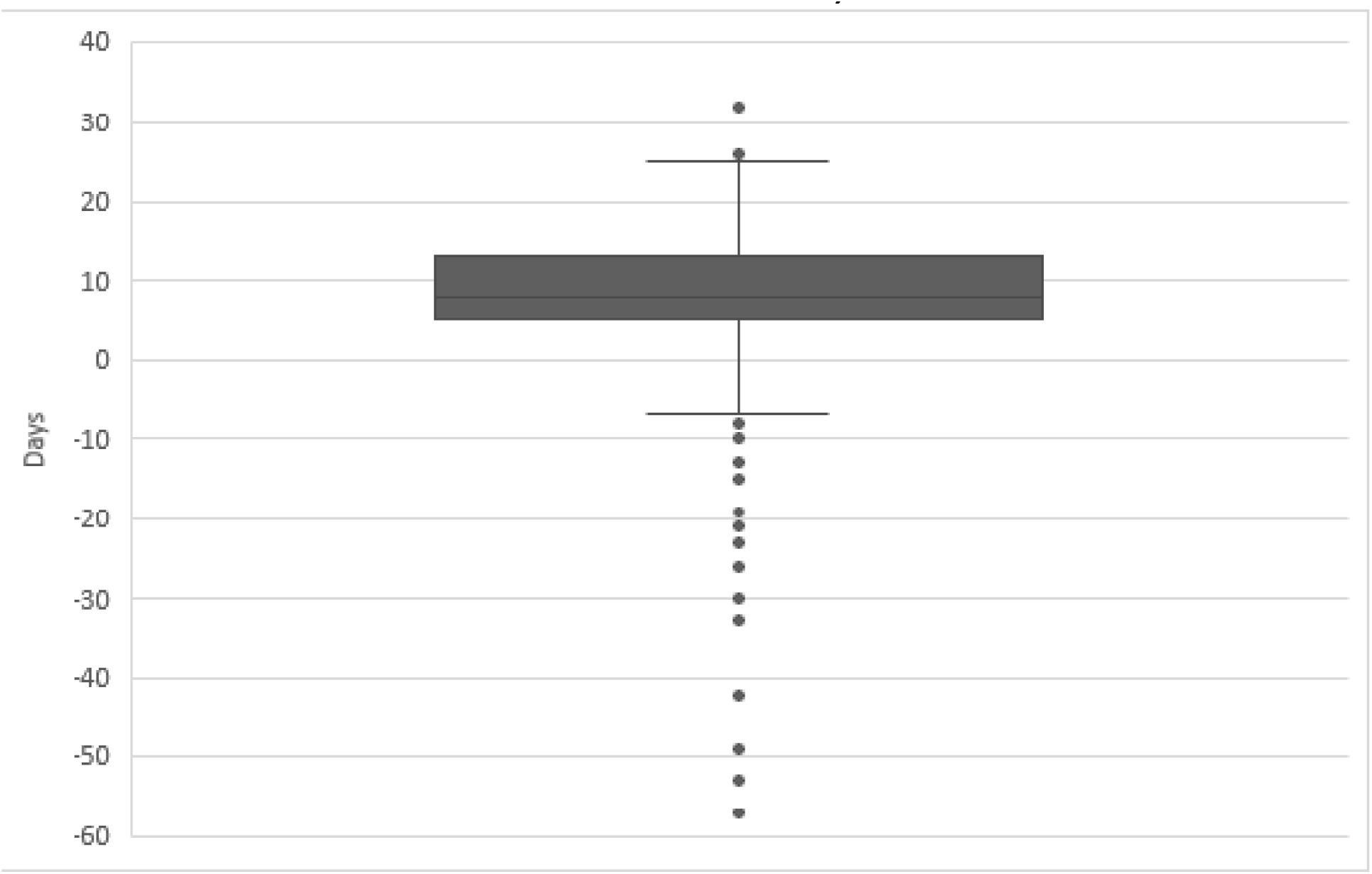
**Days from ICU admission to secondary infection specimen date on a patient level where day zero is date of ICU admission (negative values are where secondary infection onset occurred before ICU admission and positive values are where onset was after ICU admission, n=3,919).**

CFR declined most markedly in patients without COVID-19-associated infection (W1: 30.3%, n=17,221/56,815; W3: 9.2%, n=1,999/21,715) (Table 1). A smaller reduction was observed in preceding/co-infection (W1: 38.6%, n=580/1,504; W3: 26.7%, n=81/303), whereas CFR among patients with secondary infection remained static across waves (W1: 32.1%, n=568/1,767 and W3: 32.5%, n=222/684, peaked in W2: 38.6%, n=1,429/3,703) (Table 1).

## Discussion

In this large, national two-year retrospective study in England, 5.8% of hospitalised COVID-19 patients had evidence of concurrent bacterial and/or fungal infection. Preceding and co-infection were rare (0.7% and 1.5%, respectively), with the majority classified as secondary infection (3.5%). The prevalence of preceding infection remained low and stable over the three waves, peaking at 1.2% in June 2020, while co-infection varied modestly between 0.4 and 3.6% per month. Secondary infection showed temporal peaks, reaching 9.7% in February 2020 and 7.5% February 2021.

These rates are substantially lower than those reported in pandemic and seasonal influenza, where co- and secondary infections rates of 17-23% are reported [2–4]. Patients with secondary infection had higher rates of ICU admission compared to those in other groups. However, this likely reflects both increased microbiological sampling and a higher baseline risk of healthcare-associated infection (HCAI) in critically ill patients [10]. Indeed, among ICU-admitted patients, the vast majority (95.2%) of secondary infections were identified following ICU admission, consistent with invasive procedures (e.g. mechanical ventilation) and intensified sampling.

The microbiological profile observed was broadly consistent with previous English and international studies [5, 24, 25]. *E. coli* and *S. aureus* predominated in bloodstream infections across the waves among preceding/co-infections. *E. faecium* emerged as the second most common organism in secondary infections after *E. coli* during waves one and two, exceeding *S. aureus*. In LRT samples, *P. aeruginosa*, *S. aureus* and *E. coli* were the most frequently identified. In contrast, *S. pneumoniae* and *H. influenzae* infections were rarely detected, differing from earlier reports [5, 17]. This discrepancy likely reflects our restriction to invasive infection, whereas prior studies included non-invasive diagnostic modalities. Furthermore, the microbiological spectrum in COVID-19 differed from influenza, where *S. pneumoniae* is a dominant co-pathogen [3].

CFR patterns were consistent with previous observations. Patients without COVID-19-associated infection had the lowest CFR and experienced the greatest decline across waves, aligning with broad improvements in COVID-19 management [26]. In contrast, CFR in those with secondary infection remained persistently high, reinforcing the clinical impacts of HCAIs. Previous studies have reported similar findings, including higher CFRs in secondary infection compared to co-infection (26.5 versus 23.0) [5], and increased CFR associated with COVID-19-associated fungal infections in ICU settings [14].

Despite the low prevalence of bacterial and fungal infection, antibiotic prescribing was high (>70%). This likely reflects early pandemic uncertainty, where empirical antibiotic use was guided by influenza-associated co-infection patterns [2, 16, 17, 25]. Although prescribing declined over time, consistent with updated NICE guidance recommending against routine antibiotic use in the absence of suspected bacterial co-infection [27], the reduction was modest. Similar trends have been reported in national prescribing data [16]. Penicillin with beta-lactamase inhibitors were the most commonly used antibiotics, which are classified as ‘watch’ antibiotics in the UK AWaRe framework [28]. Among patients with confirmed infection, antibiotic use was comparable to previous reports, though published data remain limited and often lack granularity regarding antimicrobial class and indication [29].

This study has several limitations. First, reliance on positive microbiological samples precluded assessment of sampling practices, which may have varied over time (e.g. reducing aerosol-generating respiratory sampling). Second, vaccination status was unavailable, limiting our ability to assess its impact. Third, although patient-level prescribing data were available, information on timing, duration, and route of administration were lacking. Fourth, misclassification of infection status is possible due to restriction to bloodstream and LRT samples (in addition to confirmed CDI), the potential for blood stream isolates to reflect non-respiratory sources, and limited LRT sampling early in the pandemic. Lastly, fewer cases were included in wave 3, which may affect comparability across waves.

## Conclusion

In this large multi-centre study spanning three COVID-19 waves in England, preceding and co-infections were consistently rare, while secondary infection occurred more frequently but remained uncommon overall. Secondary infections were associated with persistently high CFR, underscoring the impact of HCAIs on patient outcomes. Despite low infection rates, antibiotic prescribing was high through the study period. These findings support antimicrobial stewardship strategies that discourage empirical antibiotic use in hospitalised COVID-19 patients in the absence of strong clinical suspicion of bacterial co-infection, in line with NICE guidance [30]. Where antibiotics are indicated, empirical regimens should reflect the predominance of *E. coli* and *S. aureus*. Finally, given the differential rates of preceding/co-infections, early diagnostic differentiation between SARS-CoV-2, influenza, and other viral pathogens should be prioritised at hospital admission.

## Data Availability

No additional data are available.

## Acknowledgements

The authors would like to thank Professor Vicki Chalker for her contributions in applying for grant funding to enable this project. We would also like to thank the HiRoSp team in UKHSA’s Immunisation and Vaccine Preventable Diseases division for providing data on invasive *H. influenzae* and *S. pneumoniae* infections. The authors would like to thank the ISARIC4C Investigators for their work collecting the data used in this study.

## Contribution

Conceptualisation: CSB, AH, SMG ; Data curation: CD, EM, NK, GB-H, AH; Formal analysis: CD, NK, GB-H, SMG; Funding acquisition: CSB, AH, SMG ; Methodology: CD, EM, LT, RH, CSB, AH, SMG; Project administration: CD, ELM; Supervision: CSB, AH, SMG; Visualisation: CD; Writing-original draft: CD, ELM, NK; Writing-review & editing: CD, EM, NK, GB-H, JE, LT, RH, CJF, TMD, MGS, JKB, CSB, AH, SMG.

## References

1. Filip, R., et al., Global Challenges to Public Health Care Systems during the COVID-19 Pandemic: A Review of Pandemic Measures and Problems. J Pers Med, 2022. 12(8).

2. Qiao, M., et al., The prevalence of influenza bacterial co-infection and its role in disease severity: A systematic review and meta-analysis. J Glob Health, 2023. 13: p. 04063.

3. Klein, E.Y., et al., The frequency of influenza and bacterial coinfection: a systematic review and meta-analysis. Influenza Other Respir Viruses, 2016. 10(5): p. 394–403.

4. Bott, H., Bacterial co-infection and antimicrobial use in hospital-attended patients with laboratory-confirmed influenza infection: a systematic review and meta-analysis. medRxiv, 2026.

5. Gerver, S.M., et al., National surveillance of bacterial and fungal coinfection and secondary infection in COVID-19 patients in England: lessons from the first wave. Clin Microbiol Infect, 2021. 27(11): p. 1658–1665.

6. Langford, B.J., et al., Bacterial co-infection and secondary infection in patients with COVID-19: a living rapid review and meta-analysis. Clin Microbiol Infect, 2020. 26(12): p. 1622–1629.

7. Gajic, I., et al., The emergence of multi-drug-resistant bacteria causing healthcare-associated infections in COVID-19 patients: a retrospective multi-centre study. J Hosp Infect, 2023. 137: p. 1–7.

8. Patton, M.J., et al., COVID-19 bacteremic co-infection is a major risk factor for mortality, ICU admission, and mechanical ventilation. Crit Care, 2023. 27(1): p. 34.

9. Wu, H.Y., et al., Coronavirus disease 2019 (COVID-19) associated bacterial coinfection: Incidence, diagnosis and treatment. J Microbiol Immunol Infect, 2022. 55(6 Pt 1): p. 985–992.

10. Garcia-Vidal, C., et al., Incidence of co-infections and superinfections in hospitalized patients with COVID-19: a retrospective cohort study. Clin Microbiol Infect, 2021. 27(1): p. 83–88.

11. Kumar, G., et al., Predictors and outcomes of healthcare-associated infections in COVID-19 patients. Int J Infect Dis, 2021. 104: p. 287–292.

12. Hughes, S., et al., Bacterial and fungal coinfection among hospitalized patients with COVID-19: a retrospective cohort study in a UK secondary-care setting. Clin Microbiol Infect, 2020. 26(10): p. 1395–1399.

13. Chong, W.H., et al., State-of-the-art review of secondary pulmonary infections in patients with COVID-19 pneumonia. Infection, 2021. 49(4): p. 591–605.

14. Borman, A.M., et al., Increased mortality in COVID-19 patients with fungal co- and secondary infections admitted to intensive care or high dependency units in NHS hospitals in England. J Infect, 2022. 84(4): p. 579–613.

15. Antimicrobial Resistance, C., Global burden of bacterial antimicrobial resistance in 2019: a systematic analysis. Lancet, 2022. 399(10325): p. 629–655.

16. Andrews, A., et al., Surveillance of Antibacterial Usage during the COVID-19 Pandemic in England, 2020. Antibiotics (Basel), 2021. 10(7).

17. Russell, C.D., et al., Co-infections, secondary infections, and antimicrobial use in patients hospitalised with COVID-19 during the first pandemic wave from the ISARIC WHO CCP-UK study: a multicentre, prospective cohort study. Lancet Microbe, 2021. 2(8): p. e354–e365.

18. Baker, M.A., et al., The Impact of Coronavirus Disease 2019 (COVID-19) on Healthcare-Associated Infections. Clin Infect Dis, 2022. 74(10): p. 1748–1754.

19. Weiner-Lastinger, L.M., et al., The impact of coronavirus disease 2019 (COVID-19) on healthcare-associated infections in 2020: A summary of data reported to the National Healthcare Safety Network. Infect Control Hosp Epidemiol, 2022. 43(1): p. 12–25.

20. Turtle, L., et al., Outcome of COVID-19 in hospitalised immunocompromised patients: An analysis of the WHO ISARIC CCP-UK prospective cohort study. PLoS Med, 2023. 20(1): p. e1004086.

21. ISARIC. Clinical Characterisation Protocol for COVID-19. 2023; Available from: https://isaric.org/research/covid-19-clinical-research-resources/clinical-characterisation-protocol-ccp/.

22. Agency, U.H.S., Laboratory reporting to UKHSA. 2025.

23. GOV.UK, Method for assigning ethnic group in the COVID-19 Health Inequalities Monitoring for England (CHIME) tool. 2023.

24. Che Yusof, R., M.N. Norhayati, and Y. Mohd Azman, Bacterial coinfection and antibiotic resistance in hospitalized COVID-19 patients: a systematic review and meta-analysis. PeerJ, 2023. 11: p. e15265.

25. Langford, B.J., et al., Antimicrobial resistance in patients with COVID-19: a systematic review and meta-analysis. Lancet Microbe, 2023. 4(3): p. e179–e191.

26. Halford, F., et al., Temporal changes to adult case fatality risk of COVID-19 after vaccination in England between May 2020 and February 2022: a national surveillance study. J R Soc Med, 2024. 117(6): p. 202–211.

27. NICE. Therapeutics for COVID-19. 2021 02/12/2025]; Available from: https://www.nice.org.uk/guidance/ng191/chapter/4-Therapeutics-for-COVID-19#antibiotics.

28. GOV.UK, UK Access, Watch, Reserve, and Other classification for antibiotics (UK-AWaRe antibiotic classification), U.H.S. Agency, Editor. 2025.

29. Al-Hadidi, S.H., et al., The Spectrum of Antibiotic Prescribing During COVID-19 Pandemic: A Systematic Literature Review. Microb Drug Resist, 2021. 27(12): p. 1705–1725.

30. NICE. Identifying and managing co-infections. 2021 02/12/2025]; Available from: https://www.nice.org.uk/guidance/ng191/chapter/6-Identifying-and-managing-co-infections.

