## Supplementary Material for "Surveillance of bacterial and fungal infections in hospitalised COVID-19 patients: an ISARIC/WHO CCP-UK and UKHSA linkage study"

Table S1: Algorithm to group multiple records from the same patient, using different combinations of patient identifiable information.

| **Order** | **NHS number (valid)** | **Date of birth** | **Patient sex** | **Patient postcode** | **Strength** |
| --- | --- | --- | --- | --- | --- |
| 1 | Y | Y |  |  | (1) Strong |
| 2 | Y |  |  |  | (2) Good |
| 3 |  | Y | Y | Y | (2) Good |

Table S2: Algorithm to group multiple records from the same patient sourced from multiple microbiological datasets, using different combinations of patient identifiable information.

| **Order** | **NHS number (valid)** | **Date of birth** | **Patient sex** | **Patient forename** | **Patient surname** | **Hospital number** | **Patient postcode** | **Strength** |
| --- | --- | --- | --- | --- | --- | --- | --- | --- |
| 1 | Y | Y | Y | Y | Y |  |  | (1) Strong |
| 2 | Y | Y | Y |  |  |  |  | (1) Strong |
| 3 | Y | Y |  |  |  |  |  | (1) Strong |
| 4 | Y |  |  |  |  |  |  | (2) Good |
| 5 |  | Y | Y | Y | Y | Y |  | (2) Good |
| 6 |  | Y | Y |  |  | Y |  | (2) Good |
| 7 |  | Y |  |  |  | Y |  | (2) Good |
| 8 | Y |  |  |  |  | Y |  | (2) Good |
| 9 |  | Y | Y | Y | Y |  |  | (3) Reasonable |
| 10 | Y |  |  | Y | Y |  |  | (3) Reasonable |
| 11 |  | Y |  | Y | Y | Y |  | (3) Reasonable |
| 12 |  |  | Y | Y | Y | Y |  | (3) Reasonable |
| 13 |  |  |  | Y | Y | Y |  | (3) Reasonable |
| 14 |  | Y | Y* | Y* | Y |  | Y | (3) Reasonable |
| 15 | Y | Y* | Y | Y | Y |  |  | (3) Reasonable |

* where the soundex of patient surname, first initial of patient forename and fuzzy date of birth were used in place of full data.
